# A deep-learning-based two-compartment predictive model (PKRNN-2CM) for vancomycin therapeutic drug monitoring

**DOI:** 10.1101/2024.01.30.24302025

**Authors:** Bingyu Mao, Ziqian Xie, Masayuki Nigo, Laila Rasmy, Degui Zhi

**Affiliations:** McWilliams School of Biomedical Informatics, The University of Texas Health Science Center at Houston, Houston, Texas, United States; Division of Infectious Diseases, Houston Methodist Hospital, Houston, Texas, United States

**Author notes:** **Corresponding author** Degui Zhi, PhD Professor McWilliams School of Biomedical Informatics The University of Texas Health Science Center at Houston 7000 Fannin St., Suite 600, Houston, TX 77030.

**Keywords:** Vancomycin, Pharmacokinetics, Compartmental Model, Recurrent Neural Network

## Abstract

**Objective:** Vancomycin is a widely used antibiotic that requires therapeutic drug monitoring (TDM) for optimized individual dosage. The deep learning-based model PKRNN-1CM has shown the advantage of leveraging time series electronic health record (EHR) data for individualized estimation of vancomycin pharmacokinetic (PK) parameters. While one-compartment (1CM) PK models are commonly used because of their simplicity and previous trough-based clinical practices for dose adjustment, the pre-deep learning literature suggests the superiority of two-compartment models (2CM). Motivated by this, we introduce a novel deep-learning-based approach, PKRNN-2CM, for vancomycin TDM.

**Methods:** PKRNN-2CM combines RNN-driven PK parameter estimation with a 2CM PK model to predict vancomycin concentration trajectories. Training on both simulated data and real-world EHR data allows for a comprehensive evaluation of its performance.

**Results:** Experiments based on simulated data highlight PKRNN-2CM’s superiority over the simpler 1CM model PKRNN-1CM (PKRNN-2CM RMSE=1.30, PKRNN-1CM RMSE=2.50). Application to real data showcases significant improvement over PKRNN-1CM (PKRNN-2CM RMSE=5.62, PKRNN-1CM RMSE=5.84, two-sample unpaired t-test p-value=0.01), with potential further gains expected with non-trough level measurements.

**Conclusion:** PKRNN-2CM is an important improvement in vancomycin TDM, demonstrating enhanced accuracy and performance compared to the PKRNN-1CM model. This deep learning model holds potential for future individualized vancomycin TDM optimization and broader application in diverse clinical scenarios.

## Introduction

Therapeutic drug monitoring (TDM) is necessary for optimizing individual dosage regimens, particularly for drugs with narrow therapeutic ranges and high pharmacokinetic (PK) variability, such as vancomycin (Avent et al., 2013; Kang & Lee, 2009). Traditional vancomycin TDM methods include trough monitoring, linear regression, population PK, and Bayesian estimation (Avent et al., 2013). The most recent national guidelines recommend individualized dosing guided by Bayesian methods for TDM, but they may not cover diverse patient populations effectively and are often unsuitable for patients with unstable clinical conditions (Narayan et al., 2021; Rybak et al., 2020). Furthermore, these models incorporate a limited number of patient-specific variables, while neglecting other factors that could enhance predictions (Narayan et al., 2021). Deep learning models, particularly recurrent neural networks (RNNs), prove to be well-suited for modeling time-series EHR data (Rasmy et al., 2018) due to their capacity to analyze sequences of time-related events. When it comes to TDM, which inherently involves irregularly sampled and noisy time-series EHR data, RNNs become a fitting choice. As an example, the PKRNN model presented by Nigo et al. (2022), is an autoregressive RNN model with a PK prediction head, which outperforms a Bayesian vancomycin TDM model. However, the prototype PKRNN model only used a one-compartment (1CM) PK model. It is unclear if multi-compartment PK models will further improve PKRNN.

Here, we extend their work, aiming to improve the performance of the PKRNN-1CM model by incorporating a two-compartment (2CM) PK prediction head within the model framework. The determination of the number of compartments in developing population PK models is important for describing the PK of drugs, as highlighted by Shingde et al. (2019). While 1CM models assume vancomycin distributes evenly throughout the body right after the infusion, multi-compartment models exhibit rapid initial distribution followed by slower elimination, offering a more realistic representation of vancomycin distribution. Shingde et al. (2019) report that vancomycin PK has been described using one, two, and three-compartment models, while most Bayesian approaches use one or two-compartment models. Their preference for 2CM models is further supported by studies indicating their superiority in higher accuracy and lower bias when predicting vancomycin concentrations (Pryka et al., 1989). 2CM models can provide more accurate predictions, especially for critically ill patients with non-steady-state kidney functions (Cheng et al., 2022; Goti et al., 2018). Additional evidence from studies like Fernandez de Gatta et al. (1996), Wu & Furlanut (1998), and Shingde et al. (2019) show the advantages of 2CM models over 1CM models.

Despite convincing evidence supporting the superiority of 2CM PK models, the 1CM model is still widely adopted in clinical practice. This preference may be attributed to simplicity and previous trough-based common practices for dose adjustment. We hypothesize that increasing the number of non-trough-level measurements of the concentration-time curve in the development of a 2CM PK model will enhance its performance, resulting in significantly improved prediction accuracy compared to a 1CM PK model. This hypothesis is grounded in the observations of Shingde et al. (2019), who noted that, for a given patient, the two curves differ the most at the peak level and become smaller the closer to the trough level. Testing this hypothesis needs a dataset rich in non-trough-level measurements, a challenge given the sparse real-world datasets. Therefore, we turn to simulation as a valuable tool, providing a controlled environment to explore different sampling strategies and thoroughly examine the impact of various factors such as the density and timing of measurements on prediction accuracy to comprehensively assess and refine the developed 2CM PK model. This study innovatively employs actual patient information in the simulation, ensuring comparability with real-world datasets, and distinguishing it from existing works using simulated data for model comparison.

This study aims to develop an RNN-based 2CM predictive model (PKRNN-2CM) for vancomycin TDM. It is anticipated that the combination of an RNN and a 2CM PK model will provide better predictions of vancomycin concentrations than the PKRNN-1CM model, especially when there is a sufficient number of observations that occur during non-trough levels.

Our study makes the following contributions: first, our novel implementation of the 2CM PK prediction head for the PK RNN model marks an important development in methodology. Second, our results show the significantly better performance of the PKRNN-2CM model for individualized vancomycin dosing with sparse and irregularly sampled real-world data. Third, through the simulation strategy combined with realistic EHR data, we systematically benchmarked the performance of different PKRNN models under diverse conditions. Finally, different simulation options not only prove the overall superiority of the 2CM model over the 1CM model but also highlight a particularly substantial performance gap, especially when dealing with non-trough-level measurements. These findings provide valuable insights into the potential of the PKRNN-2CM model for enhancing clinical decision-making for individualized vancomycin dosing.

## Statement of Significance

### Problem

Pharmacokinetic (PK) models that can take in real-world data and generate concentration trajectories accurately are desired for the clinical use of drugs that need monitoring.

### What is Already Known

Current clinical practice relies on one-compartment (1CM) PK models, despite literature suggesting the superiority of two-compartment (2CM) models. The deep learning PKRNN-1CM model is developed and showing promise, but faces limitations due to simplicity.

### What this Paper Adds

PKRNN-2CM, our novel deep-learning model combines recurrent neural networks with a 2CM PK model, outperforms PKRNN-1CM in both simulation and real-world scenarios, provides wider applicability for deep learning-based therapeutic drug monitoring (TDM).

## Methods

### Model architecture overview

As with the PKRNN-1CM model (Nigo et al., 2022), the PKRNN-2CM model is an autoregressive RNN model containing a PK model as the prediction head. The PKRNN-1CM model consists of three components that enable the vancomycin predictions to be evaluated at any time point. First, a code embedding layer is used to input information from the EHR into the PKRNN-1CM model for every time step. Next, an RNN layer is applied to predict vancomycin elimination rates and compartment volumes, and finally, based on the output from the RNN layer, a 1CM PK layer uses the PK equation, which is an ordinary differential equation (ODE), to compute the predicted vancomycin concentration. For the PKRNN-2CM model, we extend the 1CM PK model to a 2CM PK model. A schematic representation of the structure of the PKRNN-2CM model can be found in Figure 1. The input to the model is the time-series EHR data after data preprocessing, which contains categorical data, continuous data, vancomycin dose, vancomycin serum concentration measurements, and other information relevant to each patient. For every time step, the RNN layer predicts four parameters η_1_, η_2_, η_3_, and η_4_ (Lim et al., 2014) that are related to the PK parameters. After that, the PK layer based on the 2CM PK model uses an ODE to calculate the estimated PK parameters k_1_ (elimination rate for the central compartment), V_1_ (volume for the central compartment), V_2_ (volume for the peripheral compartment), and R, which is defined by k_2_/V_2_ (k_2_ is the elimination rate for the peripheral compartment), and is using the estimated PK parameters to provide the predicted vancomycin concentration as the model output.

**Figure 1.**
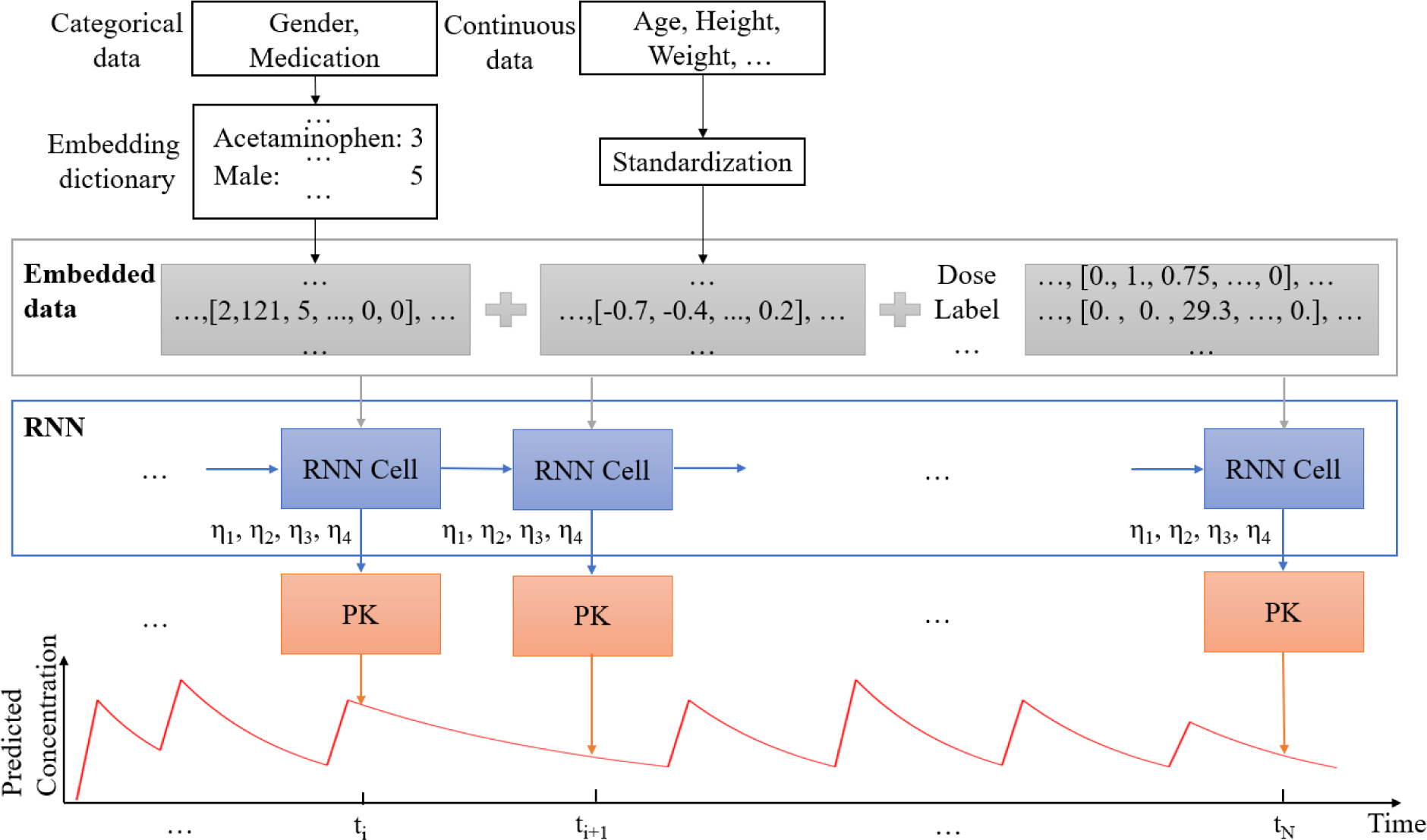
PKRNN-2CM model architecture. A schematic that shows the model structure of PKRNN-2CM by every time step (time step is defined by vancomycin administration time, vancomycin level obtained, or the end of the day). In this figure, there are N time steps. Each time step is fed with the time-series EHR data after preprocessing, which contains categorical data, continuous data, doses, measurements, and other information. After the embedding layer, the RNN layer predicts 4 parameters η_1_, η_2_, η_3_, and η_4_. The PK layer then computes the predicted vancomycin concentration based on the 2CM PK model as the model output.

PKRNN-2CM differs from PKRNN-1CM in two aspects: (1) it uses additional PK parameters and different initial values; and (2) it uses a first-order ODE system with two ODEs in order to calculate vancomycin concentrations. As Figure 2 shows, there are two PK parameters in the PKRNN-1CM model, elimination rate k and volume V, while the PKRNN-2CM model has two elimination rates, k_1_, k_2_, and two volumes, V_1_, V_2_ for the central and peripheral compartments, respectively. Furthermore, empirically we found that the result got better by removing the constraints of mass conservation as in PKRNN-1CM. Instead, in PKRNN-2CM we make the concentration continuous, which we believe is better because the model is allowed to correct its own prediction error in a smoother way.

**Figure 2.**
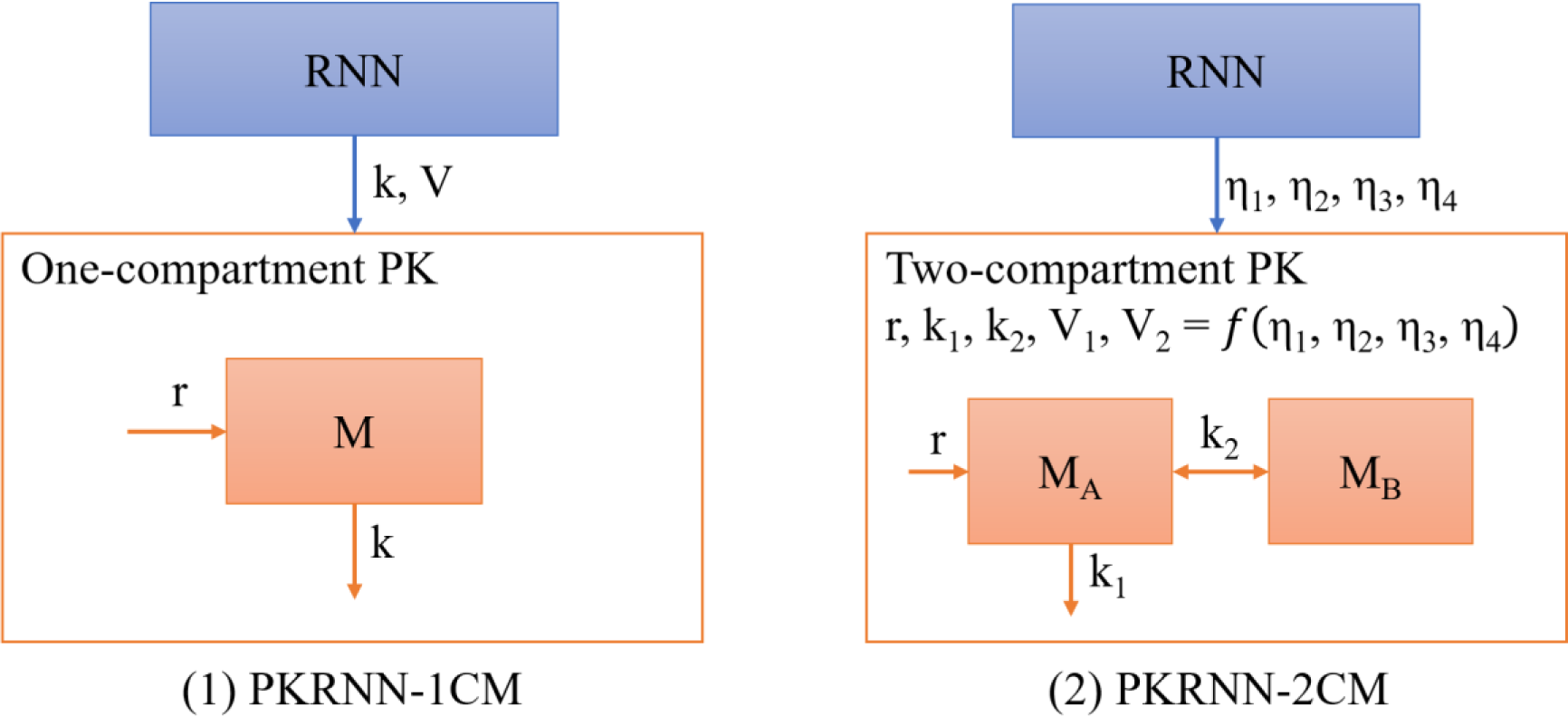
The main difference between PKRNN-1CM and PKRNN-2CM. (1) The RNN and PK layers of the PKRNN-1CM model. k: elimination rate; V: volume distribution, r: infusion rate; M: mass of the compartment. The RNN predicts k and V, where the PK layer uses these two parameters together with r and M into the ODE to calculate the vancomycin concentration. (2) The RNN and PK layers of the PKRNN-2CM model. η_1_, η_2_, η_3_, and η_4_: PK parameters that satisfy a multivariate Gaussian distribution; k_1_: elimination rate of the 2CM system; k_2_: exchange elimination rate between the two compartments; M_A_: mass of the central compartment; M_B_: mass of the peripheral compartment. The RNN predicts η_1_, η_2_, η_3_, and η_4_ to calculate 4 parameters k_1_, R, V_1_, and V_2_ for the 2CM PK model, where R=k_2_/V_2_. The PK layer uses k_1_, R, V_1_, and V_2_ as PK parameters to calculate the vancomycin concentration.

### Pharmacokinetics model details

Based on Lim et al. (2014), the four PK parameters k_1_, k_2_, V_1_, and V_2_ can be described by four related parameters η_1_, η_2_, η_3_, and η_4_ that follow a multivariate normal distribution (MVN). The RNN layer in the PKRNN-2CM predicts η_1_, η_2_, η_3_, and η_4_ where the initial values are determined according to Lim et al. (2014):

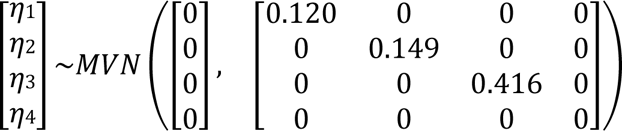

as opposed to PKRNN-1CM which uses RNN to predict k and V directly. In the PK layer, η_1_, η_2_, η_3_, and η_4_ are used to calculate PK parameters:

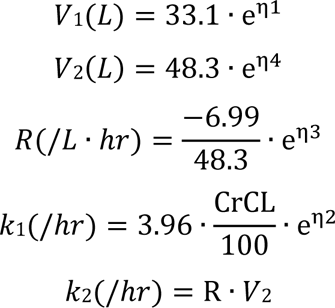

where CrCL is the creatinine clearance calculated from the Cockcroft-Gault equation:

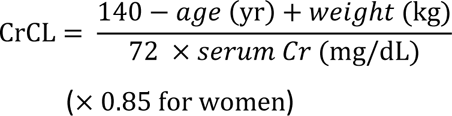

The vancomycin concentration can then be calculated using the ODE system below:

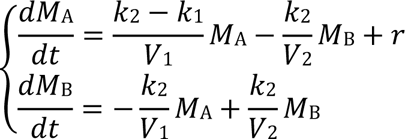

Here *M*_A_ represents the mass of the central compartment, and *M*_B_ represents the mass of the peripheral compartment. A detailed mathematical derivation can be found in the supplementary materials.

### Simulation framework

The simulation was designed to be employed as a valuable tool to guide the development and deployment of our PKRNN-2CM model by bridging the gap between the constraints of real-world data and the requirements of a comprehensive and reliable predictive system. Through this approach, we can address the limitations of sparse real-world datasets and gain deeper insights into the behavior of the PKRNN-2CM model in comparison to the PKRNN-1CM model. By generating simulated data using as much real data as possible, we bridged the gap in current works that used simulated data for model comparison. Previous studies either used one simulated standard patient to evaluate various model performances (Broeker et al., 2019) or simulated patient information for the whole patient population that did not contain any real data (Maung et al., 2022). To ensure that the simulated dataset was comparable to the original real-world dataset, the simulation was based on actual patient information such as medications and lab results. As much patient information as possible was used in the simulation to guarantee the similarity between the simulated datasets and the original real-world dataset, with the only simulated data being the measurements. The predicted vancomycin concentrations from the PKRNN-2CM model using real-world data were set as simulated measurements, and the PKRNN-2CM model that provided measurement values for simulation was defined as the underlying model. The PKRNN-1CM or PKRNN-2CM model used to test the simulated datasets was defined as an inference model.

The simulation employed in this study involves the use of vancomycin concentrations predicted by the underlying model as labels for testing the performance of the inference PKRNN-1CM and PKRNN-2CM models based on different types of simulation. The simulation framework includes some sequential steps: firstly, training a PKRNN-2CM model on real-world data to establish the underlying model for simulation. Subsequently, the predicted concentrations are utilized to generate simulated datasets. These simulated concentrations, along with other patient information, are then inputted into the inference models PKRNN-1CM or PKRNN-2CM. The final step involves evaluating the performance of the models by comparing the predicted concentration-time curve derived from the underlying model with that obtained from the inference models.

### Simulation details

The framework can be technically described into 7 steps:

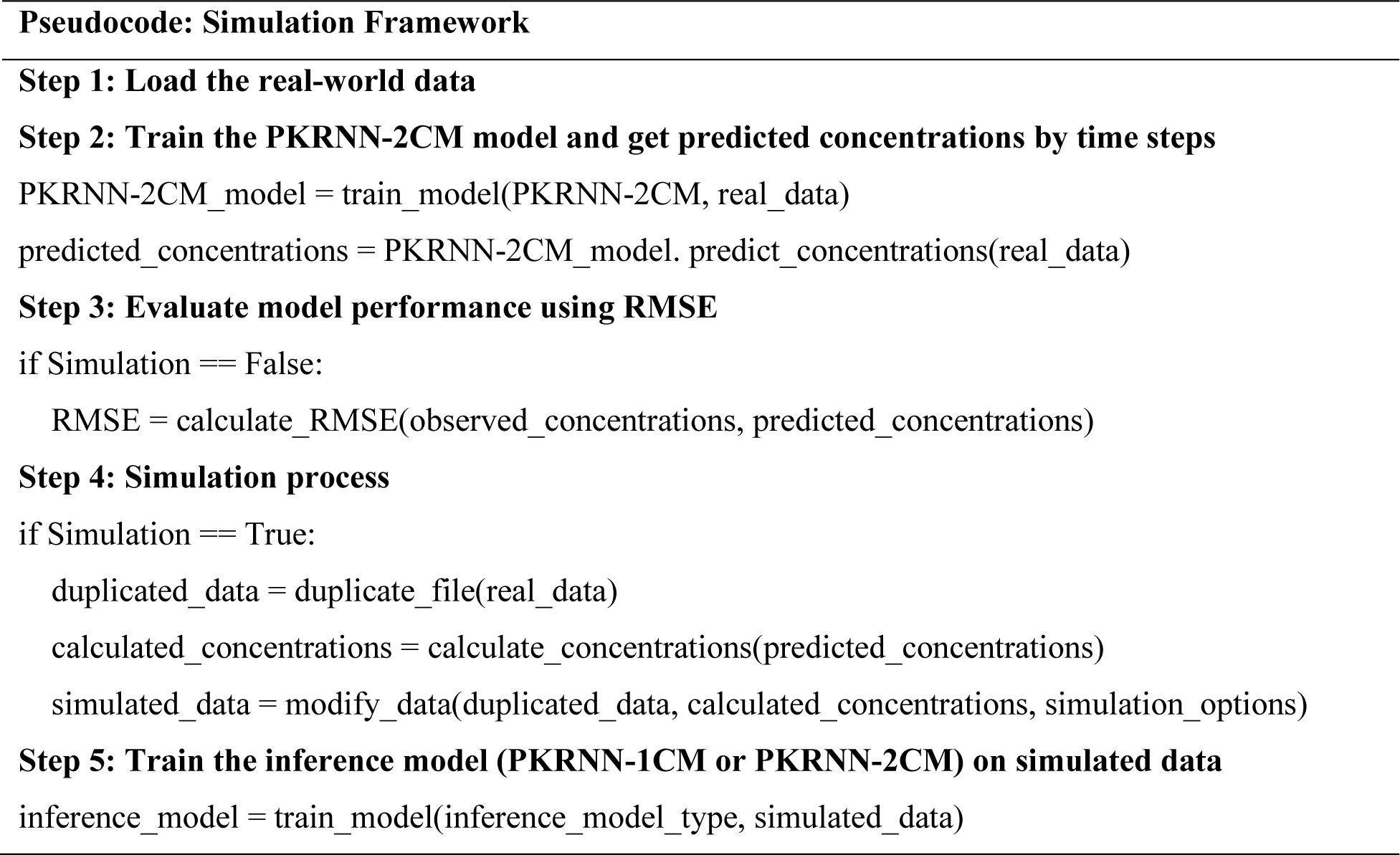
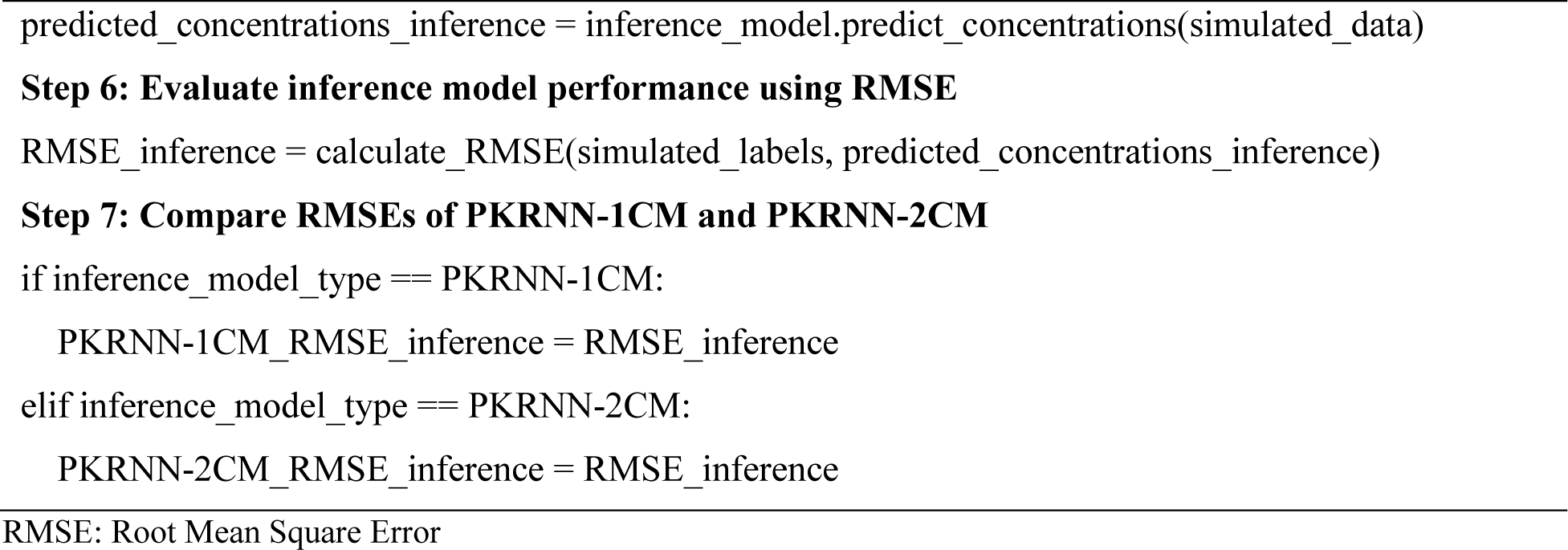

Figure 3 is a schematic diagram showing the complex simulation and evaluation process. The simulation begins with Step 1, where the real-world time series EHR data is loaded into the system. Moving forward to Step 2, the PKRNN-2CM model is trained on the real data, providing predicted concentrations based on time steps. In Step 3, when Simulation is set to False, model performance is assessed through the calculation of root mean square error (RMSE), comparing observed and predicted concentrations. Conversely, when Simulation is set to True in Step 4, the original data is duplicated, and calculated concentrations from the PKRNN-2CM model are employed to generate a simulated dataset based on simulation options. In Step 5, training the inference model (PKRNN-1CM or PKRNN-2CM) on the simulated data generated in the previous step. Step 6 evaluates the inference model’s performance using RMSE. The final step, Step 7, includes a comparative analysis of RMSEs between PKRNN-1CM and PKRNN-2CM to measure the models’ efficacy in different evaluation scenarios.

**Figure 3.**
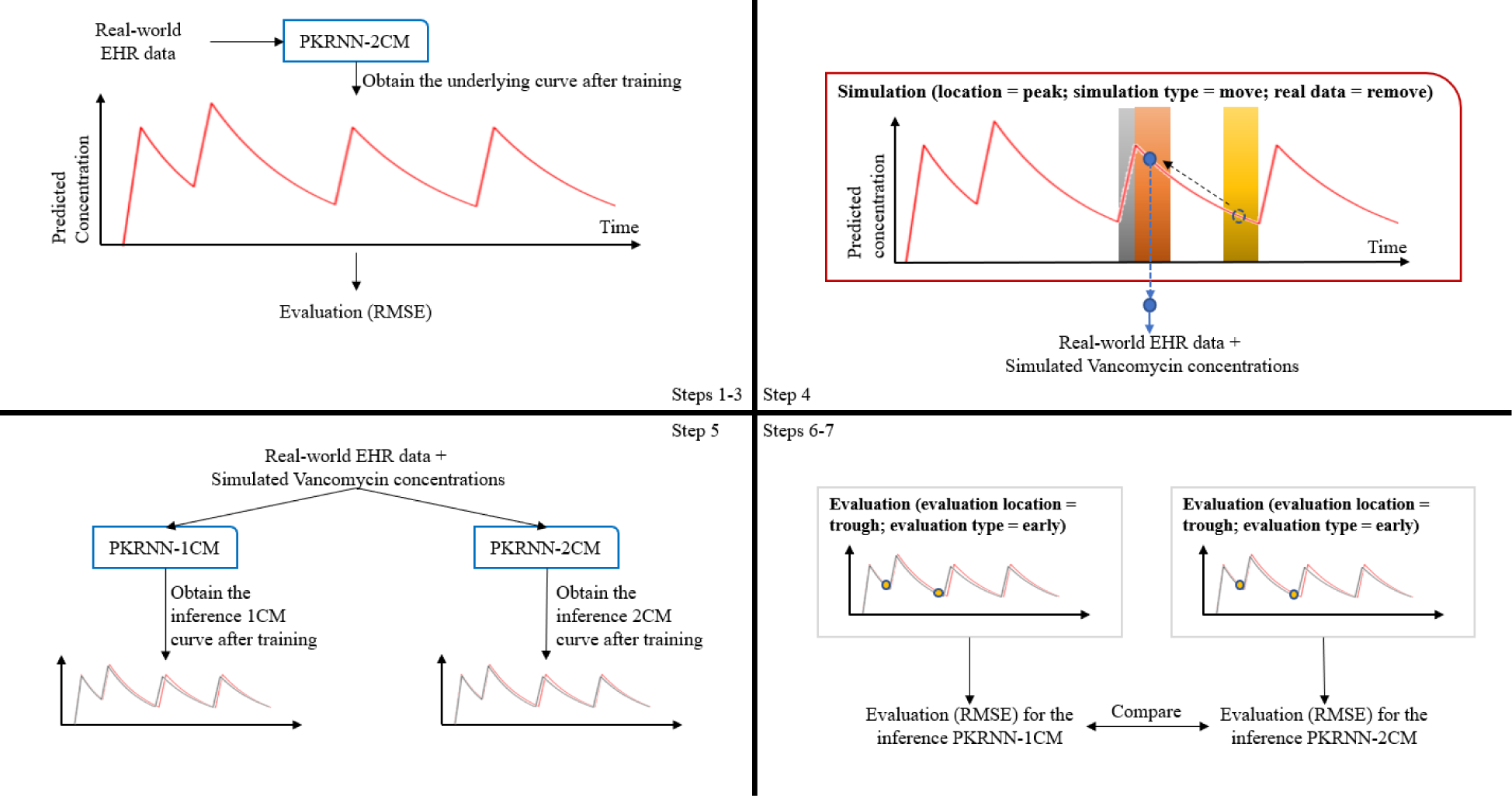
Simulation process. The top section portrays pseudocode, detailing the data loading, model training, and performance evaluation. The bottom schematic diagram visually illustrates the flow, emphasizing the sequential progression from data duplication and concentration calculation to the training and evaluation of inference models.

Figure 4 provides a comprehensive description of the simulation and evaluation process, showcasing three examples of different choices. Two simulation types were considered: moving measurements from the real-world dataset to target locations (peak or trough) or adding simulated measurements. Within the latter option, two sub-options were considered: adding measurements to the first half doses of the patient or adding measurements for every dose. Simultaneously, three location options were explored: peak, trough, and both, where the “both” option means simulating measurements at both peak and trough levels of the concentration-time curve. Addressing real data, two choices were available: to keep or remove measurements from the original real-world dataset. Evaluation criteria were also diverse, permitting assessment at peak, trough, or both locations. Moreover, there were three evaluation types provided to evaluate the early half, later half, or the whole predicted curve.

**Figure 4.**
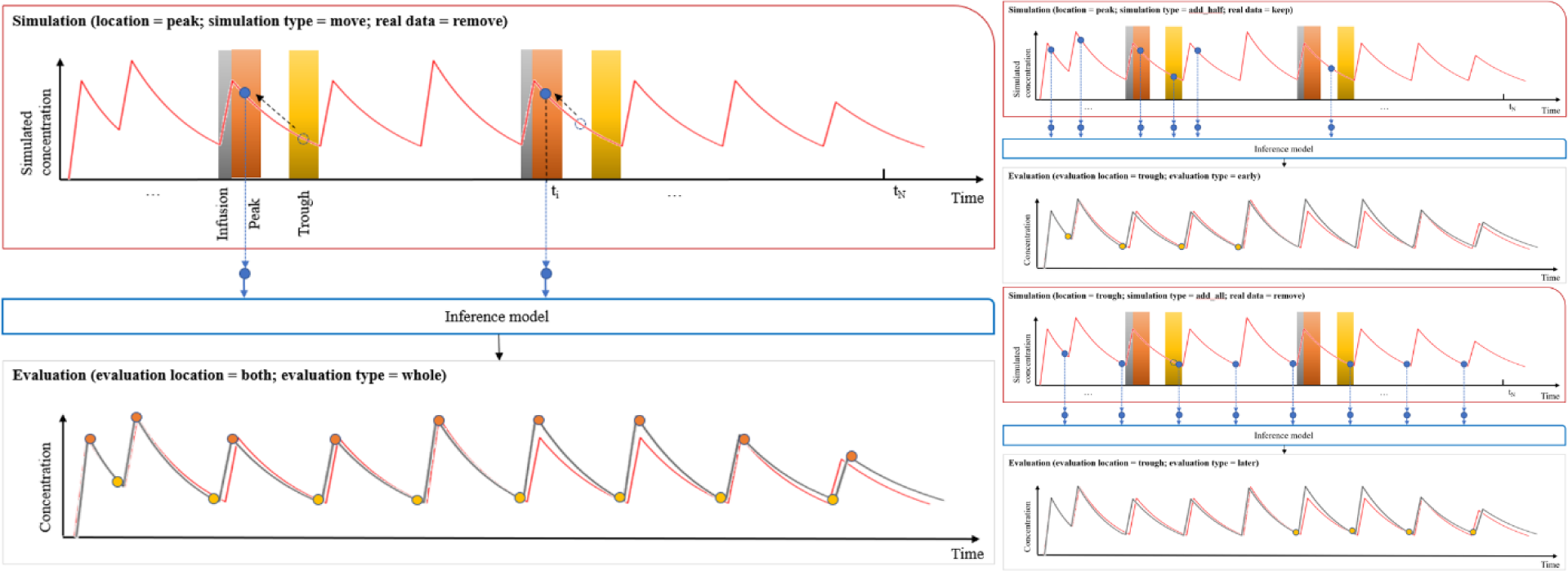
Simulation and evaluation options overview. A visual representation of the simulation and evaluation options. It showcases three examples of different choices within this framework, highlighting variations in simulation types, location options, and strategies for dealing with real data. The figure also delineates how the inference model’s predicted curve can be evaluated at peak, trough, or both locations.

### Data description

This study utilized the same dataset as Nigo et al. (2022), which was obtained from an EHR data warehouse containing information regarding encounter-level administrative data collected from Memorial Hermann Health System (MHHS), a large healthcare system based in Houston, Texas, United States. Patients over 18 who got at least one dose of vancomycin during the period from August 2019 through March 2020 were included in the study. This study excluded patients who received renal replacement therapy and patients with inappropriate timing of vancomycin levels. De-identification of the extracted cohort has been conducted in order to protect the privacy and security of the patient data. Similar to Nigo et al. (2022), our model was trained and evaluated only using encounters with at least one serum vancomycin level measured after the first vancomycin dose.

The EHR data for this study were extracted at the encounter level, including 30 laboratory tests, 5 vital signs, 324 types of medications, and demographic information. In the same manner as Nigo et al. (2022), for each encounter, the start time was determined by the earliest record time, while the end time was determined by the timestamp of the last vancomycin concentration. Vancomycin administration time, vancomycin level obtained, and the end of the day were used as time steps to update the parameters of the PKRNN-2CM model. Within the MHHS dataset, the infusion duration of vancomycin fluctuates based on the dosage: ≤1000 mg is infused over 1 hour, 1001–1500 mg over 1.5 hours, 1501–2000 mg over 2 hours, and doses exceeding 2000 mg over 2.5 hours. For model simplification, a uniform infusion rate of 1 gm/hour was adopted. It is also assumed that measurements of vancomycin should not be made during the infusion. The values that are missing are estimated with previous values based on the assumption that the clinicians did not repeat the test due to patient stability. A mean value was used when no values were measured for the patient. Z-scores are used to standardize continuous values.

### Implementation details

To establish the framework for our model implementation, we meticulously configured the code embedding and the RNN layers, and also chose training hyperparameters and optimization techniques for optimal performance. In configuring the code embedding layer, categorical data is represented by 8-dimensional vectors encompassing all medication codes. For the initialization of the embedding layer, weights were established using a Gaussian distribution. Each time step involves the input of a 48-dimensional vector (40 + 8) into the RNN layer, comprising embedded categorical data (8) and normalized continuous data (40). The RNN layer utilized a single-layer gated recurrent unit (GRU) with a hidden size of 64. The output layer is characterized by a linear layer of size (64, 4), mapping the GRU’s hidden layer to the parameters η_1_, η_2_, η_3_, and η_4_ at each time step. For model training, we employ the Adamax optimizer with a learning rate and weight decay set at 1e-1 and 0.2, respectively. The training minibatch size is configured at 50, and an early stopping mechanism, governed by the hyperparameter “patience” set to 10, is implemented to mitigate overfitting risks. The mean squared error serves as the chosen loss function for training the model. Additionally, we incorporate two regularization mechanisms. Firstly, we impose a penalty on the deviation of the predicted parameters η_1_, η_2_, η_3_, and η_4_ from the initial values in the RNN layer. The second regularization term uses the L2 norm of the first-order difference to discourage abrupt changes in the output of the RNN layer.

For the simulation implementation, several datasets were simulated from the underlying model based on different simulation options, and the sampling strategy of the simulation was based on the infusion cycle. In the simulated peak datasets, for every time step, a measurement was simulated at the next time step when a dose was present. The time intervals between these two time steps were determined based on the dose, the simulated measurements were made after 2 hours if the dose was less than or equal to 1000mg or after 3 hours if the dose was greater than 1000mg. The trough datasets simulated each measurement 1 hour before the dose. Additionally, the interval between the next time step and the simulated time step was reduced to maintain the patient’s time length unchanged.

For consistency and comparability, the PKRNN-2CM model evaluation remains the same as the PKRNN-1CM model. Depending on patient identification, the data were divided into training, test, and validation sets in a ratio of 70:15:15. A comparison of the model performance between PKRNN-2CM and PKRNN-1CM from both the original MHHS dataset and also the simulated datasets were made using RMSE. The statistical analysis to compare the PKRNN-2CM model and the PKRNN-1CM model was completed by a two-sample unpaired t-test. Additionally, the simulation evaluation was based on hours and followed the same peak and trough definitions for the sampling strategy. We ran the inference models with different simulation options to evaluate how the inference models captured the entire vancomycin concentration curve (not only the points similar to where the input measures were sampled). This study was conducted with Python 3.8 (Python Software Foundation), primarily using PyTorch 1.9.0.

A detailed code repository to reproduce our experiment can be found at: https://github.com/ZhiGroup/PK-RNN/tree/PKRNN-2CM.

## Results

This study consisted of 5,483 patients with 8,689 encounters in total. There were two parts to the descriptive analysis of the dataset, namely basic characteristics and demographics, as shown in Table 1. There was a median weight of 82.9 kg and a median height of 172cm among all patients. Age, gender, and race/ethnicity were included in the demographics data analysis. Out of 5,483 patients, 3,069 were males, which represented 55% of the population, and the median age was 61. In the patient population, 36.4% were white, which was the largest racial group when compared to less than 20% African Americans and less than 2% Asians. Moreover, 783 (14.2%) patients were Hispanic.

**Table 1.** Descriptive statistics for the study cohort.

| Characteristics | Number (%) or median (IQR) |
| --- | --- |
| ***Basic Characteristics*** |  |
| Patients | 5,483 |
| Encounters | 8,689 |
| Weight (kg) | 82.9 (65.5 – 101.6) |
| Height (cm) | 172 (165.1 – 181.1) |
| ***Demographics*** |  |
| Age | 61 (48-73) |
| Gender |  |
| Male | 3,069 (55%) |
| Race and ethnicity |  |
| White | 2,003 (36.4%) |
| African American | 1,069 (19.5%) |
| Asian | 83 (1.5%) |
| Non-Hispanic | 3,905 (71.2%) |
| Hispanic | 783 (14.2%) |

IQR: Interquartile range.

Table 2 shows that the PKRNN-2CM model exhibited improvement over the PKRNN-1CM model (the two-sample unpaired t-test p-value=1.00e-2).

**Table 2.**
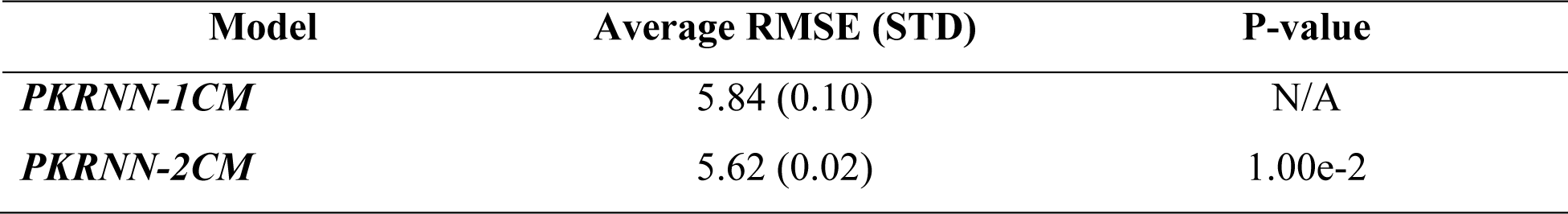
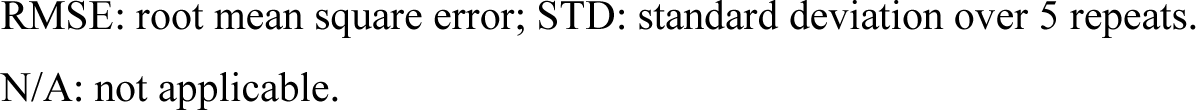
Results and comparison for PKRNN-1CM and PKRNN-2CM.

| Model | Average RMSE (STD) | P-value |
| --- | --- | --- |
| ***PKRNN-1CM*** | 5.84 (0.10) | N/A |
| ***PKRNN-2CM*** | 5.62 (0.02) | 1.00e-2 |

RMSE: root mean square error; STD: standard deviation over 5 repeats.
N/A: not applicable.

Simulation results were reported using average RMSEs from test sets. For each dataset simulated from the underlying model, we tested and compared the model performance of the inference PKRNN-1CM and the PKRNN-2CM models. Table 3 presents the summarized simulation results while additional outcomes from diverse simulation options and evaluation settings are reserved for the supplementary materials.

**Table 3.** Simulation results for simulated datasets with removing real data.

| **Add measurements**<br>**for first half doses** |  | **Average RMSE (STD)** |  | **P-value** |
| --- | --- | --- | --- | --- |
| **Simulation location** |  | **PKRNN-1CM** | **PKRNN-2CM** |  |
| Peak<sup>*</sup> |  | 2.60 (0.36) | 1.47 (0.19) | 5.07e-04 |
| Trough<sup>**</sup> |  | 3.42 (0.18) | 2.66 (0.09) | 5.20e-05 |
| Both<sup>***</sup> |  | 3.23 (0.36) | 1.59 (0.15) | 3.03e-05 |
| **Add measurements**<br>**for all doses** |  | **Average RMSE (STD)** |  | **P-value** |
| **Simulation location** |  | **PKRNN-1CM** | **PKRNN-2CM** |  |
| Peak<sup>*</sup> |  | 2.50 (0.24) | 1.30 (0.09) | 1.40e-05 |
| Trough<sup>**</sup> |  | 2.63 (0.18) | 1.53 (0.16) | 1.69e-05 |
| Both<sup>***</sup> |  | 3.04 (0.29) | 1.48 (0.15) | 1.24e-05 |

RMSE: root mean square error; STD: standard deviation over 5 repeats.
\*Peak: 2 or 3 hours post every infusion, 2 hours for infusion $\leq 1000\text{mg}$ , 3 hours for infusion $> 1000\text{mg}$ ;
\*\*Trough: 1 hour pre every infusion;
\*\*\*Both: two measurements (peak and trough) for every infusion.

Upon interpreting the results in Table 3, several noteworthy observations emerged. Firstly, simulated data consistently provided lower RMSEs than real-world data for both models, showing the regularity of the simulated datasets helps to improve the model performance. Both moving and adding measurements showed significant differences between RMSEs from the two models, particularly when adding measurements for every dose at the peak level, favoring PKRNN-2CM with more statistically significant p-values. Peak options consistently outperformed trough options for both models, emphasizing the importance of peak measurements in enhancing predictive accuracy. The strategic decision to remove real data from simulated datasets, combined with adding measurements for every dose at the peak level, resulted in the lowest RMSEs for both models, with PKRNN-2CM exhibiting superiority over PKRNN-1CM. Furthermore, PKRNN-2CM consistently demonstrated lower standard deviation across multiple runs, indicating a higher level of consistency and performance stability compared to PKRNN-1CM.

Detailed results from different evaluation options can be found in the supplementary materials, which revealed statistical significance favoring PKRNN-2CM over PKRNN-1CM. Notably, peak-level evaluations consistently gave smaller p-values than trough-level evaluations even when the simulation location was at the trough level, emphasizing the superior performance of the PKRNN-2CM model.

Figure 5 illustrates three different patient scenarios from the underlying model, and the inference models provided comparable results across these scenarios. The simulated dataset employed for inference models contained observations at the peak level after each infusion, with the option of removing real data. In Figure 5(1), the top panel showcases a patient for whom the PKRNN-2CM model, when using real data, outperformed the PKRNN-1CM model (PKRNN-1CM RMSE: 27.18; PKRNN-2CM RMSE: 21.32). Specifically, out of four observations, the PKRNN-2CM model captured three, while the PKRNN-1CM model captured none. In the bottom panel, it is evident that the inference PKRNN-2CM model captured most of the simulated observations also provide a lower RMSE (PKRNN-1CM: 17.54; PKRNN-2CM: 15.20), whereas the inference PKRNN-1CM model missed over half of them, with consistent RMSEs (PKRNN-1CM: 17.54; PKRNN-2CM: 15.20). In Figure 5(2), the PKRNN-2CM model with real data performs similarly to the PKRNN-1CM model (PKRNN-1CM RMSE: 18.28; PKRNN-2CM RMSE: 16.97). Both models missed one out of three observations, the PKRNN-1CM model caught the second observation, while the PKRNN-2CM model caught the last. The bottom panel shows that the inference PKRNN-2CM model captured simulated observations from the first dose with a lower RMSE, while the PKRNN-1CM model started capturing them nearly at the end of the curve (PKRNN-1CM RMSE: 13.24; PKRNN-2CM RMSE: 11.46). In Figure 5(3), the top panel describes a patient wherein the PKRNN-2CM model, using real data, performed less effectively than the PKRNN-1CM model (PKRNN-1CM RMSE: 14.57; PKRNN-2CM RMSE: 16.19). Out of three observations, PKRNN-1CM captured one, PKRNN-2CM captured one, and the other, missed by both models, is closer to the curve from the PKRNN-1CM model. The bottom panel indicates that the inference PKRNN-2CM model performed better on the simulated dataset (PKRNN-1CM RMSE: 11.14; PKRNN-2CM RMSE: 9.74).

**Figure 5.**
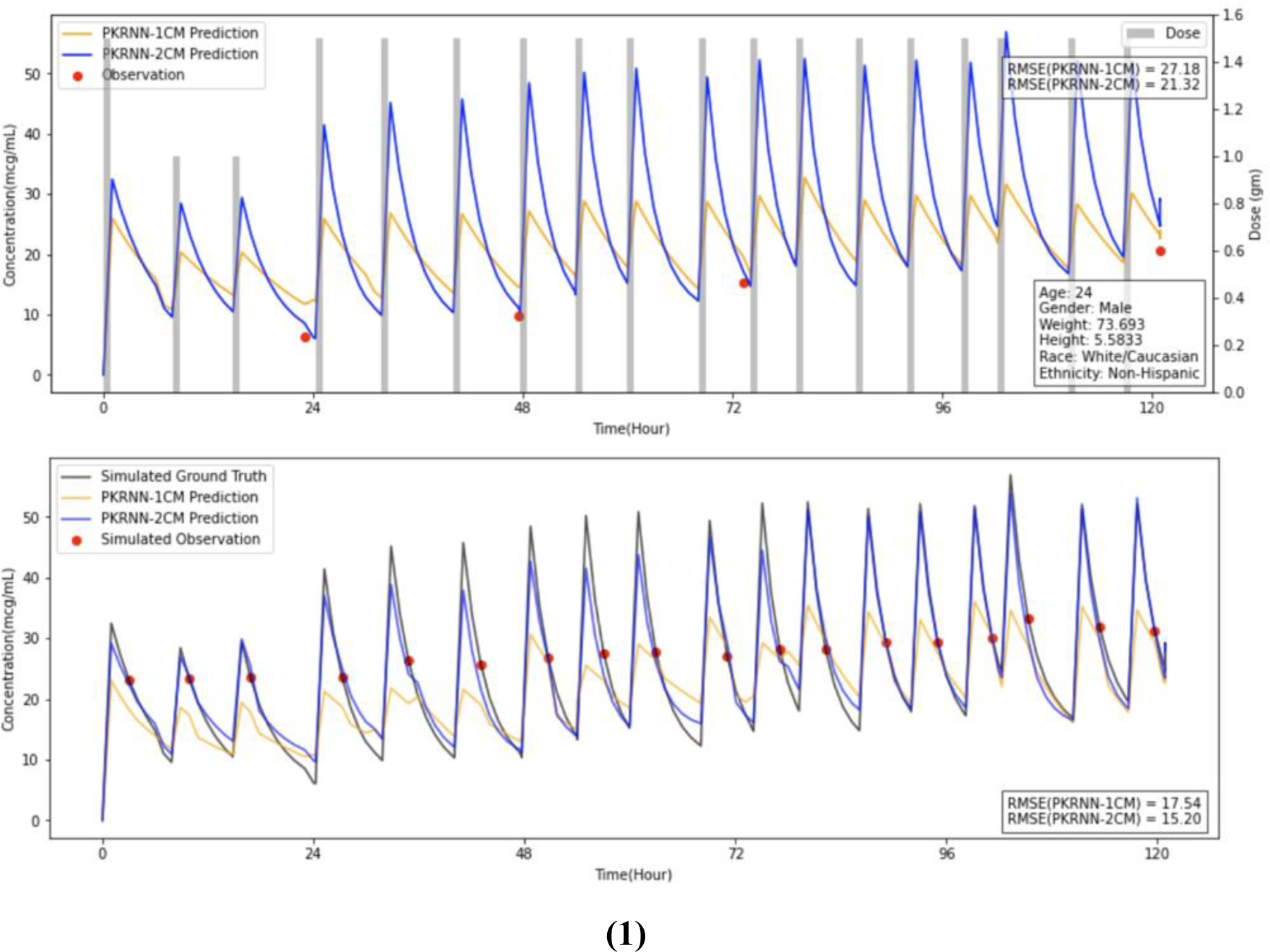
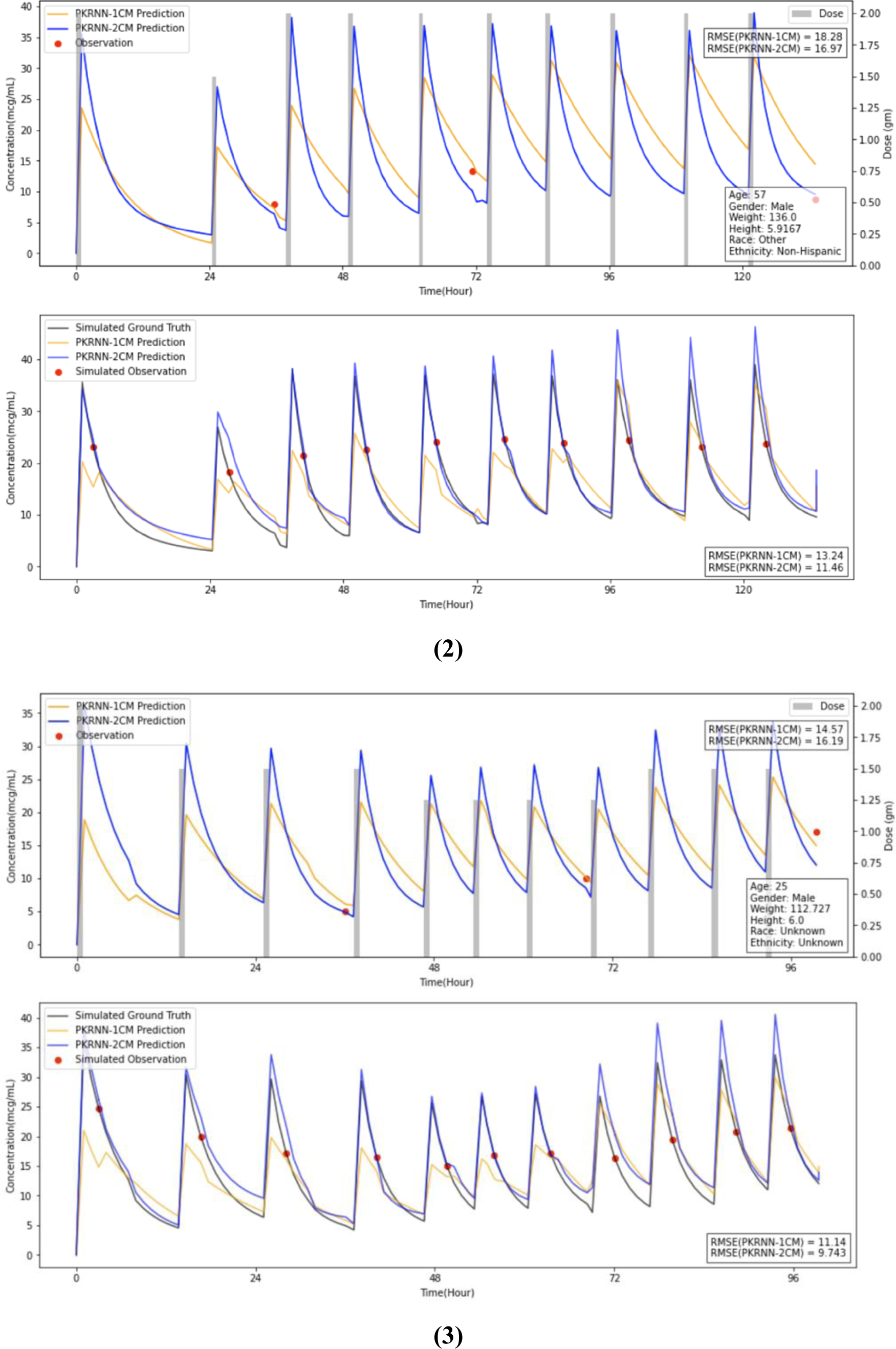
Model performance across patient scenarios. Comparison of the performance of PKRNN-1CM and PKRNN-2CM models across diverse patient scenarios. In the top panel of each subfigure, the models’ performance using real data is compared. The bottom panel assesses the models’ ability to capture simulated observations, highlighting differences in their predictive accuracy.

## Discussion

PKRNN-2CM is a novel deep-learning model that combines an RNN for vancomycin PK parameter estimation with a 2CM PK model for generating concentration trajectories, offering an innovative approach for individualized vancomycin TDM using time series EHR data. Our results demonstrate a significant improvement in the PKRNN-2CM model’s performance compared with the PKRNN-1CM model in real-world data. The results from our comprehensive simulation studies documented not only the overall superiority of the PKRNN-2CM model over the PKRNN-1CM model, but also highlighted a greater performance gap in scenarios involving non-trough-level measurements.

To the best of our knowledge, this study marks an advancement by being the first in the literature to prove the superior predictive accuracy of a 2CM PK model over the 1CM PK model using a large real-world dataset. Current experiments that compare the 1CM and 2CM PK models for prediction tasks are based either on a richly sampled small dataset (Shingde et al., 2019) or on simulated data (Broeker et al., 2019; Maung et al., 2022). While these approaches provided valuable insights, they lacked the robustness to capture the complexities in large real-world datasets with sparse measurements. In contrast, our work addresses this gap by showcasing the enhanced predictive capabilities of the 2CM PK model even under conditions of data sparsity. By validating the superiority of the PKRNN-2CM model on sparse data, our study provides a more realistic and applicable insight into the model’s performance, offering valuable implications for the clinical implementation of 2CM PK models in scenarios characterized by limited data availability. This contribution underscores the robustness of 2CM models, extending their utility to a broader range of clinical situations beyond the constraints of previous experimental settings.

To the best of our knowledge, our simulation framework is the first real-world-based simulation framework to compare the performance of 1CM and 2CM PK models. Existing studies using simulated data to compare the 1CM and 2CM PK models are either using one simulated standard patient to evaluate different model performances (Broeker et al., 2019) or simulating all the patient information for the patient population which does not contain any real data (Maung et al., 2022). The simulation in this work strives for a more realistic representation by using as much actual patient information as possible, with the only simulated components being the measurements. By grounding our simulation in real-world patient demographics, medications, and lab results, we ensure that the simulated datasets retain a level of reality to the complexities observed in actual clinical scenarios. Consequently, our simulation method bridges the gap between purely synthetic scenarios and the complex realities of clinical practice, adding credibility to our model comparisons and supporting the transferability of our findings to diverse healthcare settings.

Our simulation framework, with its diverse simulation and evaluation options, enables evaluations of PK models in real-world scenarios. The inclusion of several different options within our simulation design allows for flexibility and adaptability, making it a valuable resource for addressing various tasks related to sampling or measurement strategies. While our primary focus is on the comparative evaluation of 1CM and 2CM PK models for vancomycin prediction, the multitude of options embedded in our simulation methodology positions it as a versatile tool. This simulation framework can be used to investigate and assess different sampling or measurement strategies in distinct clinical settings and applications. In addition to different simulation options, our evaluation strategy of the simulation has a significant advantage in its ability to overcome the limitations imposed by the availability of real-world measurements. In real-world scenarios, the evaluation of PK model performance is constrained by the availability of real measurements, leaving gaps in understanding the model’s efficacy at specific levels of the concentration-time curve. However, the strategic inclusion of different evaluation options in our simulation framework circumvents these constraints, offering a comprehensive evaluation of PK model performance across various scenarios. This approach not only fills the gaps left by missing real-world measurements but also offers a forward-looking perspective for future studies. The versatility of our evaluation options opens avenues for exploring the inference model’s behavior under various conditions, enriching our understanding of its performance in a detailed manner. This adaptability positions our simulation-based evaluation as a valuable methodological advancement with broad implications for enhancing the robustness and reliability of PK model assessments.

The study conducted has some limitations that should be acknowledged. Firstly, our work explores the PKRNN-2CM model framework, a departure from the PKRNN-1CM model, introducing an additional set of parameters. In the PKRNN-1CM model, the RNN directly predicts two 1CM PK parameters. However, in the PKRNN-2CM model architecture, the RNN outputs four parameters linked to 2CM PK parameters, following an MVN (Lim et al., 2014). We did not explore the case where the RNN directly predicts 2CM PK parameters, leaving it as a potential direction for future research to compare the performance differences between the two parameterization approaches. Secondly, our simulation results suggest that peak measurements are more informative than trough measurements for model predictions, which contradicts the clinical practice of tend to use trough measurements for vancomycin TDM. We hypothesize that this paradox occurs because of the noise in real-world data that affects trough measurements more than peak measurements. Further research is needed to test this hypothesis and to explore the optimal sampling strategy for vancomycin TDM. Lastly, the PKRNN-2CM model employed in this study lacked the capacity to predict the area under the curve (AUC) of vancomycin level due to the unavailability of AUC measurements in the MHHS dataset. This poses a challenge in directly comparing our model’s performance with other existing models that use AUC as the evaluation metric.

This study has several important impacts: first, the demonstrated superior predictive accuracy of the PKRNN-2CM model over the PKRNN-1CM model, both on real-world and simulated data, signifies the ability of the PKRNN-2CM model to provide more accurate predictions from the first dose, which enables clinicians to make timely and informed dose adjustments, contributing to enhanced patient outcomes. Second, the reliance on a 2CM PK model in the PKRNN-2CM model enhances its potential for generalization to other drugs that require TDM. The more realistic PK representation in a 2CM model broadens the applicability of the PKRNN-2CM approach beyond vancomycin, laying the foundation for improved personalized TDM across various medications. Third, this study’s finding that accurate peak-level measurements exceed trough-level measurements in informativeness has implications for future clinical guidelines. While accurate peak-level measurements prove more valuable than trough-level measurements, the study recommends the concurrent use of a 2CM PK model to ensure optimal predictive accuracy, particularly for peak-level measurements. Finally, the success in developing the PKRNN-2CM model hints at a future direction for advancing model accuracy and generalizability by exploring multi-compartment models with three or more compartments. This trajectory promises further improvement in predicting the concentration-time curve for drugs that require TDM, paving the way for more realistic PK models with broader applications in clinical practice.

In conclusion, we developed the PKRNN-2CM model, which can be viewed as an improved version of the PKRNN-1CM model, and demonstrated that the 2CM PK model provides more accurate vancomycin concentration prediction than the 1CM PK model when using deep learning techniques as part of the predictive model. As far as we are aware, our PKRNN-2CM model is the first that demonstrates the 2CM PK model is significantly better than the 1CM PK model by using sparse, irregularly sampled EHR data obtained from the real world. Overall, our findings suggest that the PKRNN-2CM model has the ability to improve the accuracy of vancomycin concentration predictions and could be applied to other PK modeling tasks using time series EHR data. The results have important implications for clinical practice and highlight the potential of our PKRNN-2CM model for improving personalized vancomycin TDM.

## Supporting information

Supplementary materials (math derivation)

## Data Availability

The data used in this study contains protected health information (PHI) that is not allowed to be shared.

https://github.com/ZhiGroup/PK-RNN/tree/PKRNN-2CM

## Abbreviations

1CM: One-compartment
2CM: Two-compartment
AUC: Area Under the Curve
EHR: Electronic Health Record
GRU: Gated Recurrent Unit
MHHS: Memorial Hermann Health System
ODE: Ordinary Differential Equation
PK: Pharmacokinetic
RMSE: Root Mean Square Error
RNN: Recurrent Neural Network
TDM: Therapeutic Drug Monitoring

## Notes

### Competing Interest Statement

The authors have declared no competing interest.

### Funding Statement

This study did not receive any funding

### Author Declarations

This study was approved by the Committee for the Protection of Human Subjects in the University of Texas Health Science Center at Houston (UTHSC-H IRB) under protocol HSC-MS-19-1011.

