## Supplementary materials for "A deep-learning-based two-compartment predictive model (PKRNN-2CM) for vancomycin therapeutic drug monitoring"

#### A Additional background information

Therapeutic drug monitoring (TDM) involves measuring a specific drug's blood concentration in a patient's bloodstream at certain intervals to keep it in a safe therapeutic range, allowing individual dosage regimens to be optimized (Kang & Lee, 2009). TDM is primarily used to monitor drugs with narrow therapeutic ranges, drugs with high pharmacokinetic (PK) variability, and drugs with known serious adverse effects. Vancomycin, a widely used antibiotic as primary therapy for infections caused by methicillin-resistant Staphylococcus aureus (MRSA) since 1958, requires TDM to achieve optimal efficacy and avoid toxicity (Avent et al., 2013).

Traditionally, the PK parameters for vancomycin TDM can be estimated using four major methods, based on Avent et al. (2013): trough monitoring, linear regression, population PK methods, and Bayesian estimation. Trough concentrations, serving as a surrogate marker for the area under the concentration-time curve (AUC) to minimum inhibitory concentration (MIC) ratio, offer a simpler monitoring approach. Despite the ease of use and minimal resource requirements of trough monitoring and population PK methods, PK parameters differ greatly among models, and therefore a single vancomycin model may not be applied to diverse patient populations. While linear regression and Bayesian methods offer more precise dosage regimens to achieve targeted AUC compared to trough-based dosage regimens, they also require additional resources, such as trained healthcare professionals and improved information technology. As explained by Avent et al. (2013), Bayesian methods offer personalized vancomycin dosing recommendations by optimally utilizing both population PK model parameters and patient-specific PK data. In view of this benefit, the most recent national guidelines advocate individualized dosing guided by Bayesian methods for TDM.

It is important to note, however, that Bayesian models still have multiple limitations. As Rybak et al. (2020) point out, the Bayesian model itself is often designed for a specific group of patients and uses the characteristics of their population, therefore not covering other populations effectively. Additionally, Narayan et al. (2021) reported that these models are typically used in patients with stable PK parameters, and may not be appropriate for patients with unstable clinical conditions, such as patients in intensive care unit (ICU). Lastly, the Bayesian model often incorporates only a limited number of patient-specific variables while other related factors that could improve the prediction are not taken into account (Narayan et al., 2021).

The determination of the number of compartments in developing population PK models is important for describing the PK of drugs, as highlighted by Shingde et al. (2019). A PK compartment is a mathematical concept that refers to the space in which a drug is likely to be distributed in the body. Hence, compartment models can be used to simulate vancomycin administration, distribution, and elimination. In a 1CM PK model, the body is assumed to behave as a single, uniform compartment, which implies that vancomycin is distributed evenly throughout the body. In a multi-compartment PK model, vancomycin is initially distributed quickly to the first compartment, followed by a slower elimination phase for redistribution among other compartments. Given that the human body is inherently multi-compartmental, more compartments in the PK model tend to provide a more realistic representation of vancomycin distribution. Therefore, it is expected that more compartments should provide a more accurate description of the change in vancomycin concentration. Shingde et al. (2019) report that vancomycin PK has been described using one-, two-, and three-compartment models, while most Bayesian forecasting programs utilize one- or two-compartment models.

While historical practices and some current studies favor the simplicity of 1CM models, recent evidence suggests the superiority of two-compartment (2CM) models in many aspects. Li et al. (2021) and Kim et al. (2022) highlighted the correlation between compartment number and a more accurate predicted AUC, supporting models with more compartments. Nevertheless, Shingde et al. (2019) noted that a 2CM model is generally considered to describe vancomycin PK. Evidence from studies like Fernandez de Gatta et al. (1996), Wu & Furlanut (1998), and Shingde et al. (2019) reinforces the advantages of 2CM models over 1CM models. For example, the linear regression equations reveal significant differences in predicting maximum serum concentrations, emphasizing a 61% underestimation with the 1CM model ((Fernandez de Gatta et al., 1996).

### B A detailed flow chart for the simulation framework

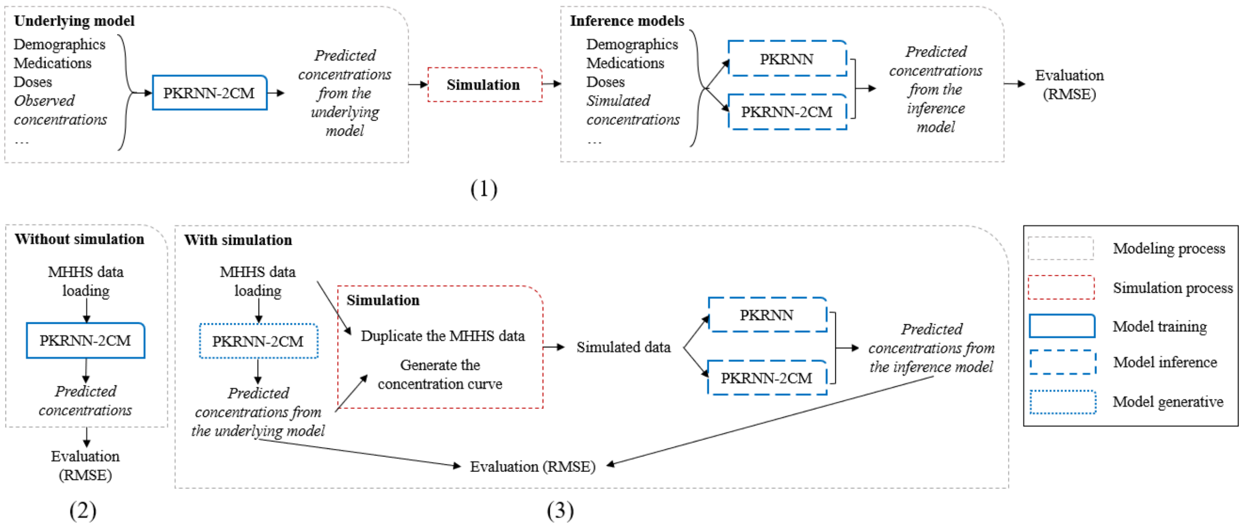


The top figure (1) shows the sequential steps: training the PKRNN-2CM model on MMHS data, generating simulated datasets, running inference models, and evaluating model performance. The bottom figures provide (2) detailed descriptions of the PKRNN-2CM training framework without simulation, (3) the simulation process.

**S Figure 1 Simulation framework**

S Figure 1 illustrates the simulation framework employed in this study. The top figure demonstrates the sequential steps involved: firstly, training a PKRNN-2CM model on MMHS data to establish the underlying model for simulation. Subsequently, the predicted concentrations are utilized to generate simulated datasets. These simulated concentrations, along with other patient information, are then inputted into the inference models PKRNN-1CM or PKRNN-2CM. The final step involves evaluating the performance of the models by comparing the predicted concentration-time curve derived from the underlying model with that obtained from the inference models. The bottom two figures provide a detailed depiction of the aforementioned framework. During model execution, there are two possible scenarios represented in the bottom figures. If the simulation parameter is set to false (bottom left), the process involves three steps: loading the MMHS data, executing the PKRNN-2CM model, and obtaining the predicted concentration. Conversely, if the simulation parameter is set to true (bottom right), the simulation commences after acquiring the predicted concentration from the underlying PKRNN-2CM model. Specifically, the MMHS data file is duplicated, and concentrations are calculated to generate the predicted concentration-time curve. The core simulation process entails utilizing the calculated concentrations to modify the previous labels and other relevant data based on different simulation options. Subsequently, the inference models are executed using the simulated data, and predicted concentrations are obtained, followed by model evaluation.

**C Model results tables based on different simulation and evaluation options**

| Test RMSE | | | Inference model | | |
|---|---|---|---|---|---|
| **Simulation type** | **Simulation location** | **Real data** | **PKRNN-1CM Avg (STD)** | **PKRNN-2CM Avg (STD)** | **P-value** |
| Move measurements | Peak | Remove | 2.97 (0.18) | 2.23 (0.12) | 1.39e-04 |
| | Trough | | 5.09 (0.17) | 4.91 (0.10) | 9.61e-02 |
| Add measurements for the first half of doses | Peak | Remove | 2.60 (0.36) | 1.47 (0.19) | 5.07e-04 |
| | Trough | | 3.42 (0.18) | 2.66 (0.09) | 5.20e-05 |
| | Both | | 3.23 (0.36) | 1.59 (0.15) | 3.03e-05 |
| Add measurements for the first half of doses | Peak | Keep | 4.05 (0.15) | 3.64 (0.04) | 6.80e-04 |
| | Trough | | 4.66 (0.14) | 4.22 (0.03) | 2.72e-04 |
| | Both | | 4.02 (0.14) | 3.21 (0.08) | 9.65e-06 |
| Add measurements for every dose | Peak | Remove | 2.50 (0.24) | 1.30 (0.09) | 1.40e-05 |
| | Trough | | 2.63 (0.18) | 1.53 (0.16) | 1.69e-05 |
| | Both | | 3.04 (0.29) | 1.48 (0.15) | 1.24e-05 |
| Add measurements for every dose | Peak | Keep | 3.16 (0.09) | 2.40 (0.03) | 1.70e-07 |
| | Trough | | 3.78 (0.13) | 3.16 (0.04) | 2.35e-05 |
| | Both | | 3.39 (0.22) | 2.04 (0.11) | 4.22e-06 |

RMSE: root mean square error; STD: standard deviation over 5 repeats.

Peak: 2 or 3 hours post every infusion, 2 hours for infusion <= 1000mg, 3 hours for infusion > 1000mg;

Trough: 1 hour pre every infusion;

Both: two measurements (peak and trough) for every infusion.

**S Table 1 Simulation results based on different simulation options**

| Test RMSE | | | Inference model | | |
|---|---|---|---|---|---|
| **Simulation location** | **Evaluation location** | **Evaluation type** | **PKRNN-1CM Avg (STD)** | **PKRNN-2CM Avg (STD)** | **P-value** |
| Peak | Peak | Early | 2.25 (0.05) | 0.60 (0.15) | 2.87e-08 |
| | | Later | 3.53 (0.21) | 0.92 (0.16) | 4.56e-08 |
| | | Whole | 4.60 (0.22) | 1.21 (0.22) | 2.02e-08 |
| | Trough | Early | 0.20 (0.01) | 0.13 (0.03) | 1.76e-03 |
| | | Later | 0.30 (0.02) | 0.21 (0.02) | 2.00e-04 |
| | | Whole | 0.40 (0.02) | 0.27 (0.04) | 3.43e-04 |
| | Both | Early | 1.79 (0.05) | 0.54 (0.13) | 1.24e-07 |
| | | Later | 3.56 (0.21) | 0.95 (0.16) | 4.48e-08 |
| | | Whole | 4.32 (0.22) | 1.20 (0.21) | 3.24e-08 |
| Trough | Peak | Early | 1.90 (0.19) | 0.73 (0.11) | 6.50e-06 |
| | | Later | 2.26 (0.22) | 0.93 (0.18) | 1.27e-05 |
| | | Whole | 3.27 (0.31) | 1.32 (0.22) | 7.33e-06 |
| | Trough | Early | 0.20 (0.02) | 0.13 (0.01) | 3.15e-05 |
| | | Later | 0.35 (0.02) | 0.24 (0.02) | 6.36e-05 |
| | | Whole | 0.45 (0.03) | 0.30 (0.02) | 3.04e-05 |
| | Both | Early | 1.58 (0.16) | 0.59 (0.11) | 7.71e-06 |
| | | Later | 2.32 (0.21) | 0.98 (0.17) | 9.14e-06 |
| | | Whole | 3.10 (0.29) | 1.25 (0.22) | 7.91e-06 |

RMSE: root mean square error; STD: standard deviation over 5 repeats.

Peak: 2 or 3 hours post every infusion, 2 hours for infusion <= 1000mg, 3 hours for infusion > 1000mg;

Trough: 1 hour pre every infusion;

Both: two measurements (peak and trough) for every infusion.

**S Table 2 Simulation results based on different evaluation options for simulation type “move” and real data “remove”**

| **Test RMSE** | | | **Inference model** | | |
|---|---|---|---|---|---|
| **Simulation location** | **Evaluation location** | **Evaluation type** | **PKRNN-1CM Avg (STD)** | **PKRNN-2CM Avg (STD)** | **P-value** |
| Peak | Peak | Early | 2.12 (0.18) | 0.80 (0.04) | 6.13e-07 |
| | | Later | 0.16 (0.02) | 0.11 (0.02) | 1.76e-03 |
| | | Whole | 0.34 (0.02) | 0.19 (0.01) | 1.45e-06 |
| | Trough | Early | 0.41 (0.03) | 0.24 (0.02) | 1.12e-05 |
| | | Later | 1.00 (0.13) | 0.33 (0.02) | 8.16e-06 |
| | | Whole | 1.40 (0.13) | 0.63 (0.04) | 2.76e-06 |
| | Both | Early | 1.92 (0.22) | 0.77 (0.04) | 6.53e-06 |
| | | Later | 2.41 (0.03) | 0.66 (0.04) | 2.27e-12 |
| | | Whole | 2.45 (0.05) | 0.84 (0.05) | 4.91e-11 |
| Trough | Peak | Early | 3.86 (0.05) | 1.20 (0.07) | 4.53e-12 |
| | | Later | 0.17 (0.00) | 0.10 (0.01) | 5.52e-08 |
| | | Whole | 0.31 (0.01) | 0.21 (0.02) | 4.93e-06 |
| | Trough | Early | 0.40 (0.01) | 0.26 (0.02) | 1.70e-06 |
| | | Later | 2.13 (0.03) | 0.56 (0.03) | 4.45e-13 |
| | | Whole | 2.50 (0.04) | 0.88 (0.05) | 2.85e-11 |
| | Both | Early | 3.70 (0.04) | 1.15 (0.06) | 2.07e-12 |
| | | Later | 1.27 (0.09) | 0.39 (0.04) | 1.16e-07 |
| | | Whole | 1.37 (0.06) | 0.66 (0.08) | 7.87e-07 |
| Both | Peak | Early | 2.08 (0.07) | 0.85 (0.09) | 2.23e-08 |
| | | Later | 0.15 (0.01) | 0.09 (0.01) | 7.54e-05 |
| | | Whole | 0.34 (0.02) | 0.19 (0.01) | 5.09e-06 |
| | Trough | Early | 0.40 (0.03) | 0.23 (0.02) | 5.55e-06 |
| | | Later | 0.90 (0.05) | 0.31 (0.04) | 1.53e-07 |

| | | | | |
|---|---|---|---|---|
| | Whole | 1.45 (0.06) | 0.71 (0.08) | 3.96e-07 |
| Both | Early | 1.87 (0.06) | 0.82 (0.09) | 4.32e-08 |
| | Later | 2.12 (0.18) | 0.80 (0.04) | 6.13e-07 |
| | Whole | 3.10 (0.29) | 1.25 (0.22) | 7.91e-06 |

RMSE: root mean square error; STD: standard deviation over 5 repeats.

Peak: 2 or 3 hours post every infusion, 2 hours for infusion <= 1000mg, 3 hours for infusion > 1000mg;

Trough: 1 hour pre every infusion;

Both: two measurements (peak and trough) for every infusion.

**S Table 3 Simulation results based on different evaluation options for simulation type "add_half" and real data "remove"**

| **Test RMSE** | | | **Inference model** | | |
|---|---|---|---|---|---|
| **Simulation location** | **Evaluation location** | **Evaluation type** | **PKRNN-1CM Avg (STD)** | **PKRNN-2CM Avg (STD)** | **P-value** |
| Peak | Peak | Early | 1.32 (0.07) | 0.32 (0.02) | 3.26e-09 |
| | | Later | 1.42 (0.12) | 0.47 (0.05) | 5.97e-07 |
| | | Whole | 2.18 (0.15) | 0.64 (0.05) | 3.95e-08 |
| | Trough | Early | 0.16 (0.01) | 0.09 (0.00) | 4.59e-06 |
| | | Later | 0.30 (0.02) | 0.18 (0.02) | 1.05e-04 |
| | | Whole | 0.37 (0.03) | 0.22 (0.02) | 3.05e-05 |
| | Both | Early | 0.99 (0.06) | 0.26 (0.02) | 1.42e-08 |
| | | Later | 1.49 (0.12) | 0.52 (0.05) | 3.24e-07 |
| | | Whole | 1.98 (0.14) | 0.63 (0.05) | 9.38e-08 |
| Trough | Peak | Early | 2.43 (0.12) | 0.63 (0.13) | 4.02e-08 |
| | | Later | 2.46 (0.12) | 0.76 (0.14) | 7.63e-08 |
| | | Whole | 3.89 (0.17) | 1.11 (0.22) | 3.98e-08 |
| | Trough | Early | 0.18 (0.02) | 0.10 (0.02) | 1.21e-04 |
| | | Later | 0.30 (0.01) | 0.19 (0.04) | 5.71e-04 |
| | | Whole | 0.39 (0.02) | 0.23 (0.05) | 2.52e-04 |
| | Both | Early | 2.14 (0.12) | 0.53 (0.12) | 7.93e-08 |
| | | Later | 2.50 (0.11) | 0.79 (0.15) | 8.40e-08 |
| | | Whole | 3.72 (0.17) | 1.06 (0.21) | 5.10e-08 |
| Both | Peak | Early | 1.34 (0.19) | 0.44 (0.06) | 1.57e-05 |
| | | Later | 1.50 (0.18) | 0.64 (0.07) | 2.27e-05 |
| | | Whole | 2.24 (0.28) | 0.87 (0.07) | 1.29e-05 |
| | Trough | Early | 0.36 (0.40) | 0.10 (0.02) | 2.31e-01 |
| | | Later | 0.31 (0.03) | 0.18 (0.02) | 1.07e-04 |

| | | | | |
|---|---|---|---|---|
| | Whole | 0.38 (0.03) | 0.22 (0.02) | 3.61e-05 |
| Both | Early | 0.98 (0.15) | 0.33 (0.05) | 3.32e-05 |
| | Later | 1.57 (0.17) | 0.68 (0.07) | 9.89e-06 |
| | Whole | 2.03 (0.25) | 0.81 (0.08) | 1.44e-05 |

RMSE: root mean square error; STD: standard deviation over 5 repeats.

Peak: 2 or 3 hours post every infusion, 2 hours for infusion <= 1000mg, 3 hours for infusion > 1000mg;

Trough: 1 hour pre every infusion;

Both: two measurements (peak and trough) for every infusion.

**S Table 4 Simulation results based on different evaluation options for simulation type "add_half" and real data "keep"**

| Test RMSE | | | Inference model | | |
| --- | --- | --- | --- | --- | --- |
| Simulation location | Evaluation location | Evaluation type | PKRNN-1CM Avg (STD) | PKRNN-2CM Avg (STD) | P-value |
| Peak | Peak | Early | 1.52 (0.09) | 0.30 (0.02) | 6.09e-09 |
| | | Later | 1.75 (0.12) | 0.33 (0.04) | 2.11e-08 |
| | | Whole | 2.57 (0.17) | 0.51 (0.04) | 1.34e-08 |
| | Trough | Early | 0.16 (0.02) | 0.07 (0.01) | 2.46e-05 |
| | | Later | 0.26 (0.02) | 0.12 (0.02) | 2.66e-06 |
| | | Whole | 0.34 (0.02) | 0.16 (0.02) | 2.41e-06 |
| | Both | Early | 1.23 (0.11) | 0.22 (0.02) | 1.11e-07 |
| | | Later | 1.79 (0.13) | 0.36 (0.04) | 2.64e-08 |
| | | Whole | 2.38 (0.20) | 0.47 (0.05) | 7.66e-08 |
| Trough | Peak | Early | 2.56 (0.15) | 0.49 (0.06) | 5.83e-09 |
| | | Later | 4.23 (0.29) | 0.78 (0.13) | 2.26e-08 |
| | | Whole | 5.36 (0.34) | 1.03 (0.16) | 1.18e-08 |
| | Trough | Early | 0.14 (0.01) | 0.06 (0.01) | 4.56e-05 |
| | | Later | 0.25 (0.03) | 0.13 (0.02) | 8.15e-05 |
| | | Whole | 0.30 (0.03) | 0.15 (0.03) | 3.95e-05 |
| | Both | Early | 2.28 (0.13) | 0.38 (0.04) | 2.41e-09 |
| | | Later | 4.25 (0.29) | 0.80 (0.13) | 2.06e-08 |
| | | Whole | 5.20 (0.32) | 0.96 (0.15) | 9.44e-09 |
| Both | Peak | Early | 1.35 (0.07) | 0.41 (0.05) | 3.07e-08 |
| | | Later | 1.63 (0.11) | 0.44 (0.03) | 3.30e-08 |
| | | Whole | 2.31 (0.13) | 0.67 (0.06) | 1.56e-08 |
| | Trough | Early | 0.13 (0.01) | 0.08 (0.03) | 9.44e-03 |
| | | Later | 0.26 (0.03) | 0.16 (0.03) | 1.49e-03 |

| | | | | |
|---|---|---|---|---|
| | Whole | 0.32 (0.03) | 0.19 (0.05) | 2.06e-03 |
| Both | Early | 1.01 (0.07) | 0.30 (0.05) | 1.58e-07 |
| | Later | 1.67 (0.11) | 0.49 (0.03) | 3.40e-08 |
| | Whole | 2.09 (0.13) | 0.62 (0.06) | 2.89e-08 |

RMSE: root mean square error; STD: standard deviation over 5 repeats.

Peak: 2 or 3 hours post every infusion, 2 hours for infusion <= 1000mg, 3 hours for infusion > 1000mg;

Trough: 1 hour pre every infusion;

Both: two measurements (peak and trough) for every infusion.

**S Table 5 Simulation results based on different evaluation options for simulation type “add_all” and real data “remove”**

| **Test RMSE** | | | **Inference model** | | |
|---|---|---|---|---|---|
| **Simulation location** | **Evaluation location** | **Evaluation type** | **PKRNN-1CM Avg (STD)** | **PKRNN-2CM Avg (STD)** | **P-value** |
| Peak | Peak | Early | 1.63 (0.10) | 0.29 (0.02) | 6.47e-09 |
| | | Later | 1.86 (0.09) | 0.35 (0.05) | 2.64e-09 |
| | | Whole | 2.75 (0.14) | 0.52 (0.05) | 1.34e-09 |
| | Trough | Early | 0.15 (0.01) | 0.08 (0.01) | 2.10e-06 |
| | | Later | 0.27 (0.02) | 0.14 (0.01) | 4.84e-06 |
| | | Whole | 0.34 (0.02) | 0.17 (0.01) | 2.06e-06 |
| | Both | Early | 1.33 (0.08) | 0.24 (0.02) | 5.96e-09 |
| | | Later | 1.91 (0.09) | 0.39 (0.05) | 2.24e-09 |
| | | Whole | 2.57 (0.12) | 0.50 (0.06) | 1.43e-09 |
| Trough | Peak | Early | 2.66 (0.13) | 0.52 (0.06) | 2.15e-09 |
| | | Later | 4.37 (0.32) | 0.84 (0.05) | 1.96e-08 |
| | | Whole | 5.55 (0.35) | 1.11 (0.08) | 8.13e-09 |
| | Trough | Early | 0.15 (0.02) | 0.07 (0.01) | 1.07e-04 |
| | | Later | 0.27 (0.03) | 0.15 (0.01) | 5.67e-05 |
| | | Whole | 0.33 (0.03) | 0.18 (0.02) | 5.24e-05 |
| | Both | Early | 2.39 (0.11) | 0.43 (0.06) | 1.62e-09 |
| | | Later | 4.39 (0.31) | 0.86 (0.06) | 1.75e-08 |
| | | Whole | 5.39 (0.34) | 1.05 (0.08) | 7.28e-09 |
| Both | Peak | Early | 1.40 (0.06) | 0.43 (0.04) | 3.42e-09 |
| | | Later | 1.70 (0.1) | 0.43 (0.04) | 1.50e-08 |
| | | Whole | 2.4 (0.12) | 0.69 (0.05) | 3.90e-09 |
| | Trough | Early | 0.13 (0.00) | 0.08 (0.02) | 3.60e-03 |
| | | Later | 0.26 (0.02) | 0.16 (0.05) | 3.49e-03 |

| | | | | |
|---|---|---|---|---|
| | Whole | 0.31 (0.02) | 0.20 (0.05) | 3.43e-03 |
| Both | Early | 1.05 (0.07) | 0.29 (0.05) | 1.41e-07 |
| | Later | 1.75 (0.10) | 0.48 (0.06) | 1.87e-08 |
| | Whole | 2.17 (0.12) | 0.61 (0.08) | 2.07e-08 |

RMSE: root mean square error; STD: standard deviation over 5 repeats.

Peak: 2 or 3 hours post every infusion, 2 hours for infusion <= 1000mg, 3 hours for infusion > 1000mg;

Trough: 1 hour pre every infusion;

Both: two measurements (peak and trough) for every infusion.

**S Table 6 Simulation results based on different evaluation options for simulation type “add_all” and real data “keep”**
