## Supplementary materials (math derivation) for "A deep-learning-based two-compartment predictive model (PKRNN-2CM) for vancomycin therapeutic drug monitoring"

### D Mathematical derivation

The mathematical derivation of the two-compartment pharmacokinetic (PK) model can be described by the following ordinary differential equation (ODE) system:

$$\begin{cases} \frac{\mathrm{d}A}{\mathrm{d}t} = -\frac{A}{V_1}k_1 + (\frac{A}{V_1} - \frac{B}{V_2})k_2 + r = \frac{k_2 - k_1}{V_1}A - \frac{k_2}{V_2}B + r \\ \frac{\mathrm{d}B}{\mathrm{d}t} = -(\frac{A}{V_1} - \frac{B}{V_2})k_2 = -\frac{k_2}{V_1}A + \frac{k_2}{V_2}B \end{cases} \tag{1}$$

Here, $A$ and $B$ represent the masses of the central and peripheral compartments, respectively. The volumes of the central and peripheral compartments are denoted by $V_1$ and $V_2$ respectively. The parameters $k_1$ and $k_2$ represent the elimination rates through the central and peripheral compartments, while $r$ corresponds to the infusion rate of the system.
To simplify the representation, we rewrite the ODE system in matrix form:

$$\frac{\mathrm{d}}{\mathrm{d}t}\begin{pmatrix} A \\ B \\ 1 \end{pmatrix} = \begin{pmatrix} \frac{k_2-k_1}{V_1} & -\frac{k_2}{V_2} & r \\ -\frac{k_2}{V_1} & \frac{k_2}{V_2} & 0 \\ 0 & 0 & 0 \end{pmatrix} \begin{pmatrix} A \\ B \\ 1 \end{pmatrix}$$

To simplify the derivation, we introduce a parameter $R = \frac{k_2}{V_2}$, assuming that $k_2$ and $V_2$ decrease at the same rate when transitioning from the two-compartment system to a one-compartment system. We denote the matrix above as $M$, and with this parameter $R$, the updated matrix $M$ becomes:

$$\begin{pmatrix} \frac{k_2-k_1}{V_1} & -R & r \\ -\frac{k_2}{V_1} & R & 0 \\ 0 & 0 & 0 \end{pmatrix}$$

Next, we diagonalize the matrix $M$ by calculating its eigenvectors and eigenvalues. After diagonalization, we obtain $M = S \cdot J \cdot S^{-1}$ where $S$ is a matrix containing the eigenvectors, and $J$ is a diagonal matrix with the eigenvalues on the diagonal:

$$S = \begin{pmatrix} \frac{rV_1}{k_1} & \frac{k_1-k_2+RV_1+\sqrt{(-k_1+k_2+RV_1)^2+4k_1RV_1}}{2k_2} & \frac{k_1-k_2+RV_1-\sqrt{(-k_1+k_2+RV_1)^2+4k_1RV_1}}{2k_2} \\ \frac{rk_2}{k_1R} & 1 & 1 \\ 1 & 0 & 0 \end{pmatrix}$$

$$J = \begin{pmatrix} 0 & 0 & 0 \\ 0 & \frac{-k_1+k_2+RV_1-\sqrt{(-k_1+k_2+RV_1)^2+4k_1RV_1}}{2k_2} & 0 \\ 0 & 0 & \frac{-k_1+k_2+RV_1+\sqrt{(-k_1+k_2+RV_1)^2+4k_1RV_1}}{2k_2} \end{pmatrix}$$

$S^{-1} =$

$$\begin{pmatrix} 0 & 0 & 1 \\ \frac{k_2}{\sqrt{(-k_1+k_2+RV_1)^2+4k_1RV_1}} & \frac{-k_1+k_2-RV_1+\sqrt{(-k_1+k_2+RV_1)^2+4k_1RV_1}}{2\sqrt{(-k_1+k_2+RV_1)^2+4k_1RV_1}} & -\frac{k_2r(-k_1+k_2+RV_1+\sqrt{(-k_1+k_2+RV_1)^2+4k_1RV_1})}{2k_1R\sqrt{(-k_1+k_2+RV_1)^2+4k_1RV_1}} \\ -\frac{k_2}{\sqrt{(-k_1+k_2+RV_1)^2+4k_1RV_1}} & \frac{k_1-k_2+RV_1+\sqrt{(-k_1+k_2+RV_1)^2+4k_1RV_1}}{2\sqrt{(-k_1+k_2+RV_1)^2+4k_1RV_1}} & -\frac{k_2r(k_1-k_2-RV_1+\sqrt{(-k_1+k_2+RV_1)^2+4k_1RV_1})}{2k_1R\sqrt{(-k_1+k_2+RV_1)^2+4k_1RV_1}} \end{pmatrix}$$

The eigenvalues $\lambda_1$ and $\lambda_2$ are given by:

$$\lambda_1 = \frac{-k_1 + k_2 + RV_1 - \sqrt{(k_1 - k_2 - RV_1)^2 + 4k_1RV_1}}{2V_1}$$
$$\lambda_2 = \frac{-k_1 + k_2 + RV_1 + \sqrt{(k_1 - k_2 - RV_1)^2 + 4k_1RV_1}}{2V_1} \tag{2}$$

To express the ODE system in terms of the transformed variables, we rewrite it as $\frac{\mathrm{d}X}{\mathrm{d}t} = M \cdot X$, and obtain $\frac{\mathrm{d}S^{-1}X}{\mathrm{d}t} = S^{-1} \cdot M \cdot X = J \cdot S^{-1} \cdot X$ after diagonalization, which implies this linear transformation turns $X$ to $S^{-1}X$. We define $C$ and $D$ as linear combinations of $A$, $B$, and $r$ after transformation, resulting in:

$$\sqrt{(k_1 - k_2 - RV_1)^2 + 4k_1RV_1}\frac{\mathrm{d}C}{\mathrm{d}t} = k_2\frac{\mathrm{d}A}{\mathrm{d}t} + (\lambda_2 - R)V_1\frac{\mathrm{d}B}{\mathrm{d}t} - \frac{rV_1k_2}{Rk_1}\lambda_2$$
$$\sqrt{(k_1 - k_2 - RV_1)^2 + 4k_1RV_1}\frac{\mathrm{d}D}{\mathrm{d}t} = -k_2\frac{\mathrm{d}A}{\mathrm{d}t} - (\lambda_1 - R)V_1\frac{\mathrm{d}B}{\mathrm{d}t} + \frac{rV_1k_2}{Rk_1}\lambda_1$$

By defining $\delta = \sqrt{(k_1 - k_2 - RV_1)^2 + 4k_1RV_1}$, we can combine the previous equations to obtain:

$$\delta(\frac{\mathrm{d}C}{\mathrm{d}t}(\lambda_1 - R) + \frac{\mathrm{d}D}{\mathrm{d}t}(\lambda_2 - R)) = (\lambda_1 - \lambda_2)k_2(\frac{\mathrm{d}A}{\mathrm{d}t} + \frac{rV_1}{Rk_1}) \tag{3}$$

When the initial values are set to $A(0) = B(0) = 0$, the following equation holds:

$$\begin{cases} \delta C(t) = -\frac{rV_1k_2}{Rk_1}\lambda_2 e^{\lambda_1 t} \\ \delta D(t) = \frac{rV_1k_2}{Rk_1}\lambda_1 e^{\lambda_2 t} \end{cases} \tag{4}$$

Furthermore, based on the eigenvalue definition, we can derive $\lambda_1\lambda_2 = -\frac{Rk_1}{V_1}$ and $\lambda_2 - \lambda_1 = \frac{\delta}{V_1}$. By combining (3) and (4), we obtain the following equation:

$$A(t) = \frac{(\lambda_1V_2 + k2)V_1}{k_2V_2}C(t) + \frac{(\lambda_2V_2 + k2)V_1}{k_2V_2}D(t) + rV_1$$
$$== \frac{r(\lambda_1 - R)}{\lambda_1(\lambda_2 - \lambda_1)}(1 - e^{\lambda_1 t}) - \frac{r(\lambda_2 - R)}{\lambda_2(\lambda_2 - \lambda_1)}(1 - e^{\lambda_2 t}) \tag{5}$$

For subsequent time steps, let us assume that for time step $t_i$, the initial values are $A_{t_{i-1}} = X_{i-1}$ and $B_{t_{i-1}} = Y_{i-1}$. Consequently, we can derive the following equation:

$$\delta C_{t_{i-1}} = (k_2X_{i-1} + (\lambda_2 - R)V_1Y_{i-1} - \frac{rV_1k_2}{Rk_1}\lambda_2)e^{\lambda_1 t} + \frac{rV_1k_2}{Rk_1}\lambda_2 e^{\lambda_1(t-d)}$$
$$\delta D_{t_{i-1}} = (-k_2X_{i-1} - (\lambda_1 - R)V_1Y_{i-1} + \frac{rV_1k_2}{Rk_1}\lambda_1)e^{\lambda_2 t} - \frac{rV_1k_2}{Rk_1}\lambda_1 e^{\lambda_2(t-d)}$$

We set the vancomycin dose as $d_{t_i} = s_i$, allowing us to derive:

$$
\begin{aligned}
A_{t_i} &= \frac{\delta}{V_1}(C_{t_i}\frac{(\lambda_1 - R)V_1}{k_2(\lambda_1 - \lambda_2)} + D_{t_i}\frac{(\lambda_2 - R)V_1}{k_2(\lambda_1 - \lambda_2)}) - \frac{rk_2}{V_1\lambda_1\lambda_2} \\
&= \frac{\lambda_1 - R}{\lambda_2 - \lambda_1}e^{2\lambda_1\Delta t_i}(X_{i-1} - \frac{(\lambda_2 - R)V_1}{k_2}Y_{i-1}) + \frac{\lambda_2 - R}{\lambda_2 - \lambda_1}2e^{\lambda_2\Delta t_i}(\frac{(\lambda_1 - R)V_1}{k_2}Y_{i-1} - X_{i-1}) \\
&+ \frac{r}{\lambda_1\lambda_2(\lambda_2 - \lambda_1)}(\lambda_2(\lambda_1 - R)(1 + e^{\lambda_1\Delta t_i})(e^{\lambda_1(\Delta t_i - s_i)} - e^{\lambda_1\Delta t_i}) \\
&+ \lambda_1(\lambda_2 - R)(e^{\lambda_2\Delta t_i} - 1)(e^{\lambda_2(\Delta t_i - s_i)} - e^{\lambda_2\Delta t_i})) \qquad (6)
\end{aligned}
$$

By following this procedure, we can determine the vancomycin concentration in the central compartment at the $i$-th time step.
